# Late reproduction is associated with extended female survival but not with familial longevity

**DOI:** 10.1101/2023.06.26.23291896

**Authors:** Annelien C de Kat, Femke Roelofs, P. Eline Slagboom, Frank JM Broekmans, Marian Beekman, Niels van den Berg

## Abstract

**Objective:** Female reproductive health comprises adequate oocyte quality and quantity, normal fecundability, a normal course of pregnancy, and the delivery of healthy offspring. General aging patterns and the maintenance of somatic health have been associated with female reproductive health. However, it is unknown whether better long-term somatic maintenance is directly related to reproductive outcomes, and whether there is a shared genetic predisposition underlying both somatic and reproductive aging. Here we investigate whether reproductive health is related to female lifespan or familial longevity.

**Design:** Observational study

**Subjects:** 10,255 female members of a multigenerational historical cohort (1812-1910), the LINKing System for historical family reconstruction (LINKS) and 1,258 females from 651 long-lived families in the Leiden Longevity Study.

**Main outcome measures:** The association between reproductive characteristics and longevity was studied both on an individual and familial level. Individual survival was studied in relation to age at last childbirth and total number of children. Familial longevity was studied through parental survival and related to age at last childbirth, total number of children and a polygenic risk score (PRS) for age at menopause.

**Results:** Females giving birth to their last child at a higher age lived longer: for each year increase in the age of the birth of the last child, a woman’s lifespan increased with 0.06 years (22 days) (p<0.005). Females who lived to be in the top 10% survivors of their birth cohort (n=2,241, 21.9%) on average gave birth to their last child at a 1% later age than the remaining cohort (IRR=1.01, p<0.005).

Females with 1 or 2 long-lived parents did not have a higher mean age of last childbirth. There was neither a significant association between an increasing number of long-lived ancestral family members (familial longevity), nor the PRS.

**Conclusion:** Female reproductive health associates with a longer lifespan and with survival to more extreme ages (longevity). The heritable component in familial longevity, however, does not associate to extended reproductive health and the PRS underlying age at menopause does not explain familial longevity. Other factors in somatic maintenance that support a longer lifespan are likely to have an impact on reproductive health.

## Introduction

Female reproductive health encompasses the evolution of being born with a complete set of oocytes, fertility and pregnancy, to the deterioration of ovarian quality and quantity, and ultimately postmenopausal health. It is widely accepted that these milestones and transitions do not stand alone but may be subject to the same processes that govern overall somatic aging and health ^1^. This relationship has not yet been fully clarified and it thus remains unknown to what extent the maintenance of somatic health is primarily essential to reproductive health or vice versa, and whether there is a genetic predisposition underlying both healthier somatic and reproductive aging.

Over the past decades a plethora of studies has sought to determine and explain the relationship between ovarian and overall somatic aging. Though there remains some dispute, several studies have observed that mothers who give birth to a child at an advanced age have a longer post-reproductive survival ^2, 3^. Studies also suggested a familial or genetic component underlying both an increasing somatic lifespan and a longer reproductive period or reproductive lifespan^4, 5^, although others have proposed a trade-off mechanism for an increasing lifespan and childbearing^6^. The latter results originate from studies with varying sample sizes and potential biases in the selection of their study population and await confirmation from well-defined, large-scale cohorts. If longevity and late reproductive aging coincide in families, the study of both traits in longevity families may reveal shared genetic loci predisposing to better maintenance of somatic and reproductive cell functions.

Genetic studies into menopause generated polygenic risk scores (PRS) that indicate risk of early menopause and involved loci in DNA repair processes, known as hallmark mechanisms of ageing ^7^. It is unknown whether the genetic component in age at menopause associates with that in familial longevity.

The current study addresses the relationship between somatic and reproductive aging and health in both a large multigenerational historical cohort and a cohort of long-living families. We aim to test whether reproductive health is associated with longevity, as well as whether members of exceptionally long-living families exhibit more favorable reproductive outcomes, compared to families from the general population. Firstly, we investigate whether lifespan (age at death) and longevity (survival to extreme ages) associate with age at last childbirth and total number of children. Secondly, we investigate whether age at last childbirth and number of children are associated with the number of long-lived parents (0, 1 or 2 long-lived parents), as an indicator of familial longevity^12^. Thirdly, we investigate whether an increasing number of long-lived ancestors associates with a PRS for age at menopause, capturing alleles associated with age at menopause, as a proxy for the total reproductive lifespan.

## Methods

### LINKS Study population

We used data from the LINKing System for historical family reconstruction (LINKS) which is a historical cohort of inhabitants of the province of Zeeland, the Netherlands, from the early 19^th^ century. The LINKS database contains demographic and genealogical information derived from the Netherlands linked vital event registration. In the Netherlands, birth, marriage and death certificates were registered from the year 1812 onward. Currently LINKS Zeeland contains 739,453 birth, 387,102 marriage, and 641,216 death certificates that were linked together to reconstruct intergenerational pedigrees and individual life courses^8^.

Two generations were identified in the dataset (Suppl Figure 1); F0 and F1, of which the F1 generation is the index generation comprising the study participants. The F0 generation was selected by identifying couples who were married between 1812 and 1850 and had at least two children, ensuring that the F1 persons had at least one sibling. The families were mutually exclusive, meaning that a parent in the F0 generation could only contribute data for a single family. From the F1 generation, the LINKS research persons (RP) were selected using the following criteria: members of the female sex, with an age of death above 50 years and a single spouse who lived until the RP was at least 50 years, and who delivered at least one child, ensuring high data quality. This selection made it possible to study the reproductive outcomes in the study population throughout the entire fertile lifespan. In both generations, a distinction was made between persons who belonged to the top 10% of survivors in their birth cohort, and those who did not. This calculation was based on Dutch lifetables, nationally collected data of survival of historic cohorts from Statistics Netherlands (CBS)^8^. Reproductive characteristics of the RPs are derived using information of their children (F3 generation). The reproductive characteristics that could reliably be extracted from the historic data were age at last childbirth and total number of children.

### Leiden Longevity Study population

The Leiden Longevity Study (LLS) was initiated in 2002 to study the mechanisms that lead to exceptional survival in good health. The LLS currently consists of 651 three-generational families, defined by siblings who have the same parents. Inclusion took place between 2002 and 2006 and initially started with the recruitment of living nonagenarian sibling pairs of European descent (F2 generation). Within a sibling pair, males were invited to participate if they were 89 years or older and females if they were 91 years or older (N=944, mean age=93 years), representing <0.5% of the Dutch population in 2001^9, 10^. Inclusion was subsequently extended to the offspring of the sibling pairs and the partners of these offspring (F3 generation). This study focuses on all F3 generation females, which are henceforth denoted as LLS RPs (offspring and partners combined).

The LLS DNA samples were genotyped using Illumina Infinium HD Human660W-Quad and OmniExpress BeadChips (Illumina, San Diego, CA, USA). DNA genotyping for LLS was performed at baseline as described in detail in Beekman et al., 2006^11^ with the Illumina Human660W and Illumina OmniExpress arrays (Illumina, San Diego, CA, USA). Genotype imputation was performed using 288,635 SNPS with SNP-wise call rate (>95%), minor allele frequency (>1%) and no derivation from the Hardy–Weinberg equilibrium (p-value > 1×10^−4^) at the Michigan Imputation Server (https://imputationserver.sph.umich.edu/index.html) with Haplotype Reference Consortium (HRC) reference panels (HRC1.1).

Mortality information was verified by birth or marriage certificates and passports whenever possible. Additionally, verification took place via personal cards which were obtained from the Dutch Central Bureau of Genealogy. In January 2021 all mortality information was updated through the Personal Records Database (PRD) which is managed by Dutch governmental service for identity information. https://www.government.nl/topics/personal-data/personal-records-database-brp. The combination of officially documented information provides very reliable and complete ancestral as well as current mortality information.

### Construction of LRC score in LLS data

Familial longevity was quantified with the Longevity Relatives Count (LRC) score. The LRC score can be interpreted as a weighted proportion (ranging between 0 and 10) ^12^. For example, an LRC score of 5 for an RP indicates 50% long-lived family members, weighted by the genetic distance between RPs and their family members.

### Construction of PRS of age at menopause in LLS data

The genome-wide association analysis for age at menopause resulted in a polygenic risk score (PRS) ^7^ that could be constructed from 290 SNPs. After we removed all T/A SNPs, SNPSs with MAF<0.01 and HWE <10^-4^, and an imputation quality <0.8 ^13^, we were able to use 195 SNPs to construct the PRS for age at menopause in the LLS data set.

### Statistical analysis

First, in LINKS, lifespan (age at death) was regressed on 1) age at last child birth (quantitative and categorical) and 2) number of children (quantitative and categorical), while adjusting for maternal birth cohort and age at marriage using a linear mixed-model with a random effect for the unique sibship ID to account for within sibship correlation.

Second, in LINKS, 1) age at last child birth and 2) number of children were regressed on a) the number of long-lived parents (0, 1 or 2 parents that belonged to the top 10% survivors of their birth cohort), as an indicator of familial longevity ^14^, and b) whether the RP survived to the top 10% of her birth cohort, while adjusting for maternal birth cohort and age at marriage using a Poisson mixed-model with a random effect for the unique sibship ID to account for within sibship correlation.

Third, in the LLS, the standardized (mean=0, SD=1) PRS of age at menopause of RPs was regressed on the number of long-lived family members, as measured with the LRC score, while adjusting for standardized birth year using a linear mixed-model with a random effect for the family ID to account for within family correlation.

All analyses were performed with R version 4.0.2.

## Results

### Study populations

In the LINKS data, 10,255 female Research Persons (RPs; the F1 generation) were identified. Collectively, they had 7,664 mothers and 7,636 fathers (the F0 generation) and 72,895 children (the F2 generation). In total, there were 7,721 unique families, taking into account that the RP generation included siblings. Mean age at death of the RPs was 73.9 (± 10.4) years and their mean number of children was 7.1 (± 3.9). Further descriptive characteristics of the RP group are described in detail in Table 1.

**Table 1.** LINKS study population selected for females that gave birth to at least one child.

| | Total number | Mean ± SD | Range |
| --- | --- | --- | --- |
| Number F1 RPs (N, % female) | 10,255 (100) | - | - |
| Top 10% survivors of their birth cohort | 2,241 (21.8) |  |  |
| Number of unique sibships (N) | 7,721 | - | - |
| Birth year (Mean) | - | 1839 | 1812-1873 |
| Age at death in years (Mean ± SD) | - | 73.9 ± 10.4 | 50-104 |
| Number of children (Mean ± SD) | 72,985 | 7.1 ± 3.9 | 1-24 |
| Number of children ≤ 4 (N, %) | 2,990 (29.2) | 2.6 ± 1.1 | 1-4 |
| Number of children ≥ 10 (N, %) | 2,771 (27.1) | 12.2 ± 2.1 | 10-24 |
| Age at first child in years (Mean ± SD) | - | 26.9 ± 4.9 | 15-49 |
| Age at last child in years (Mean ± SD) | - | 39.2 ± 5.0 | 18-51 |
| Last childbirth ≤ 40 years (N, %) | 5,190 (50.6) | 35.5 ± 4.4 | 18-40 |
| Last childbirth ≥ 45 years (N, %) | 981 (9.6) | 45.8 ± 1.0 | 45-51 |
| Age at marriage<sup>α</sup> in years (Mean ± SD) | - | 25.9 ± 4.6 | 16-46 |
| Number of identified F0 parents (N, %) | 15,300 (99.1) | - | - |
| RPs with 0 long-lived<sup>#</sup> parents (N, %) | 8,293 (80.9) | - | - |
| RPs with 1 long-lived<sup>#</sup> parent (N, %) | 1,849 (18.0) | - | - |
| RPs with 2 long-lived<sup>#</sup> parents (N, %) | 113 (1.1) | - | - |

<sup>$\alpha$</sup> In the case of multiple marriages, age at first marriage was considered. <sup>#</sup>Belonging to the top 10% survivors of their birth cohort. 15.3% (2,344) of the identified parents have a missing age at death.

The LLS cohort included 1,258 females with a mean age of 59 years.

### Females giving birth to their last child at a higher age live longer

To confirm in the LINKS dataset that mothers who give birth to a child at an advanced age have a longer post-reproductive survival, we investigated the relation between age at last child and lifespan using linear mixed-model regression analysis. We observed that for each year increase in the age of the birth of the last child, a woman has a 0.06 years (22 days) longer lifespan (p-value=2.16·10^-3^). When we compared the lifespan of females with a low (≤40 years) vs high (≥45 years) age at last child birth, we observed that females who delivered their last child beyond the age of 45 lived 1.41 years (17 months) longer than females who had their last child before 40 years (p-value=9.07·10^-5^), while adjusting for age at marriage and the mother’s birth year (Table 2).

**Table 2.** Age at last child birth and number of offspring associates with female lifespan.

|  | **N (mean/proportion)** | **Beta (CI)** | **P-value** |
| --- | --- | --- | --- |
| Age at last child (quantitative) | 10,255 (39.23) | 0.06 (0.02- 0.10) | 2.16 x·10<sup>-3</sup> |
| Age at last child (qualitative) |  |  |  |
| Group 0: ≤ 40 years | 5,190 (50.6) | REF | REF |
| Group 1: ≥ 45 years | 981 (9.6) | 1.41 (0.71-2012) | 9.07 x·10<sup>-5</sup> |
| Number of children (quantitative) | 10,255 (7.12) | 0.05 (-0.01-0.11) | 7.63 x·10<sup>-2</sup> |
| Number of children (qualitative) |  |  |  |
| Group 0: ≤ 4 | 2,990 (29.2) | REF | REF |
| Group 1: ≥ 10 | 2,771 (27.1) | 0.61 (-0.03-1.25) | 6.25 x·10<sup>-2</sup> |

All analyses are adjusted for the maternal birth year and RPs age at marriage. In addition, our research design accounts for survival of RPs and their partner up to the age of 50 years and the number of marriages. Analyses are done using mixed-model linear regression using the lme4 and lmerTest package in R. Confidence intervals have been calculated in R with the "confint" function using the Wald method. 4 separate analyses have been done with age at death as outcome; 1 where age at last child (quantitative definition) was independent variable of interest, 1 where age at last child (qualitative definition) was independent variable of interest, 1 where number of children (quantitative definition) was independent variable of interest, 1 where number of children (qualitative definition) was independent variable of interest.

The number of children may relate both to age at latest childbirth and to survival of the mother, so we investigated whether the number of children associates with female lifespan. We performed a linear mixed-model regression analysis with lifespan as outcome and the number of children as the independent variable of interest, while adjusting for age at marriage and maternal birth year. We observed that with each additional child the lifespan of females increased with 0.05 years (p-value=7.63·10^-2^). Moreover, females who had 10 or more children (N=2,771) in comparison to females who had 4 and fewer children (N=2,990) lived on average 0.61 years (18 days) longer, but this did not reach statistical significance (Table 2; p-value=6.25·10^-2^).

Next, we studied whether females who lived as long as the top 10% survivors of their birth cohort (long-lived), gave birth to their last child at higher ages than other females using mixed-model Poisson regression analysis. While adjusting for age at marriage and the RP’s maternal birth year, we observed that the top 10% survivor (N = 2,241, 21.9%)), on average gave birth to their last child at a 1% later age than the remaining cohort (N= 8,014; 78,1%) (IRR=1.01; p-value=2.75·10^-3^). Thus, with an increasing age of last child, an RP has a very small increasing chance to become long-lived. Moreover, females who lived as long as the top 10% of their birth cohort, had on average 2% more children than other females (IRR=1.02, p-value=9.19·10^-2^), although this effect was not statistically significant. We conclude from the LINKS cohort that women with more offspring and a high age at last reproduction have a longer lifespan and that top 10% female survivors of their birth cohort are capable to reproduce longer than other women.

### Females with long-lived parents do not differ from females from the general population with respect to their age at last child and number of children

High quality somatic maintenance in individuals may beneficially affect both reproductive health and longevity. It has also been suggested that familial longevity and especially heritable mechanisms drive such joint beneficial effects. To study whether familial longevity associates with reproductive health, we investigated whether females from long-lived families have more offspring and higher age at last reproduction. We observed that RPs with 1 or 2 long-lived parents do not have a significantly higher number of children (IRR-1 LL parent=0.98, p-value=1.62·10^-1^; IRR-2 LL parents=0.98, p-value=6.44·10^-1^) nor a higher mean age at last child birth (IRR-1 LL parent=0.99, p-value=5.28·10^-2^; IRR2 LL parents=1.00, p-value=9.99·10^-1^) compared to RPs without any long-lived parents (Table 3).

**Table 3.**
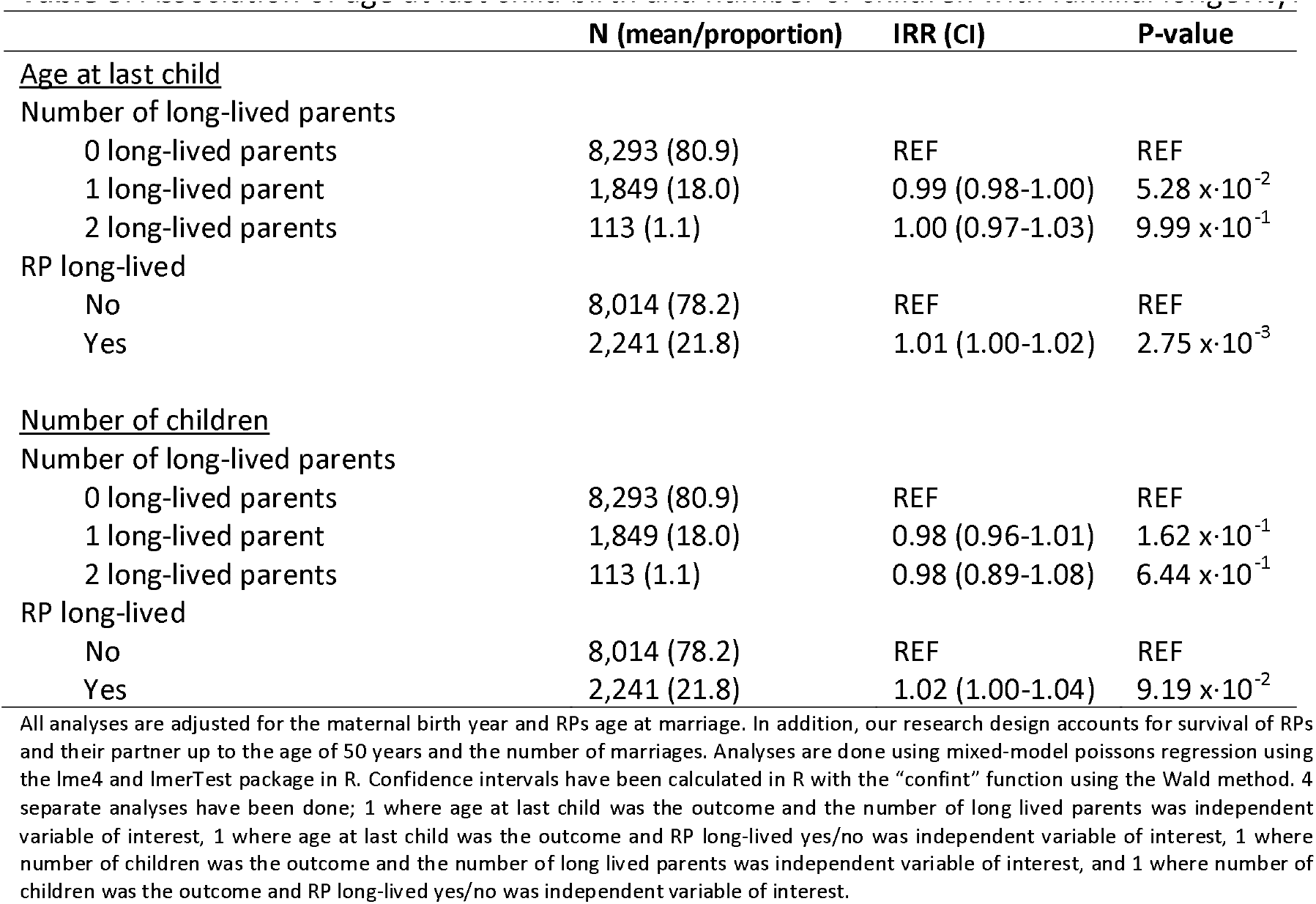
Association of age at last child birth and number of children with familial longevity.

|  | N (mean/proportion) | IRR (CI) | P-value |
| --- | --- | --- | --- |
| <u>Age at last child</u> |  |  |  |
| Number of long-lived parents |  |  |  |
| 0 long-lived parents | 8,293 (80.9) | REF | REF |
| 1 long-lived parent | 1,849 (18.0) | 0.99 (0.98-1.00) | 5.28 x·10<sup>-2</sup> |
| 2 long-lived parents | 113 (1.1) | 1.00 (0.97-1.03) | 9.99 x·10<sup>-1</sup> |
| RP long-lived |  |  |  |
| No | 8,014 (78.2) | REF | REF |
| Yes | 2,241 (21.8) | 1.01 (1.00-1.02) | 2.75 x·10<sup>-3</sup> |
| <u>Number of children</u> |  |  |  |
| Number of long-lived parents |  |  |  |
| 0 long-lived parents | 8,293 (80.9) | REF | REF |
| 1 long-lived parent | 1,849 (18.0) | 0.98 (0.96-1.01) | 1.62 x·10<sup>-1</sup> |
| 2 long-lived parents | 113 (1.1) | 0.98 (0.89-1.08) | 6.44 x·10<sup>-1</sup> |
| RP long-lived |  |  |  |
| No | 8,014 (78.2) | REF | REF |
| Yes | 2,241 (21.8) | 1.02 (1.00-1.04) | 9.19 x·10<sup>-2</sup> |

All analyses are adjusted for the maternal birth year and RPs age at marriage. In addition, our research design accounts for survival of RPs and their partner up to the age of 50 years and the number of marriages. Analyses are done using mixed-model poisson's regression using the lme4 and lmerTest package in R. Confidence intervals have been calculated in R with the "confint" function using the Wald method. 4 separate analyses have been done; 1 where age at last child was the outcome and the number of long lived parents was independent variable of interest, 1 where age at last child was the outcome and RP long-lived yes/no was independent variable of interest, 1 where number of children was the outcome and the number of long lived parents was independent variable of interest, and 1 where number of children was the outcome and RP long-lived yes/no was independent variable of interest.

### Females with long-lived parents have a similar genetic predisposition for a late menopause as other females

To further study the genetic component underlying both longevity and reproductive health we focused on the Leiden Longevity Study. We investigated whether an increasing number of long-lived ancestors of female RPs, as measured with the LRC score, associates with an increasing number of alleles marking a later age at menopause. In the Leiden Longevity Study we included a generation of RP women (F3 generation; N=1,258) with a quantified family history of longevity, as well as their genotypes through single nucleotide polymorphism (SNP) data. We calculated a PRS for age at menopause based on the most recent Genome Wide Association Study (GWAS) ^7^.

Using mixed-model linear regression, no statistically significant association between an increasing number of long-lived family members (familial longevity) and our polygenic score for age at menopause was observed (Beta=0.014 (95% CI=-0.01-0.04), P-value=2.63·10^-1^). Still, the effect size was in the expected direction: with every 10 percent increase in long-lived family members, the age at menopause, as expressed by our polygenic score, was 0.014 standard deviation lower.

## Discussion

In the historic (1812-1910) multigenerational LINKS database of Zeeland (NL), we observed that females who gave birth to their last child at a higher age, and with increasing numbers of offspring had a longer lifespan. Moreover, females who were among the top 10% survivors of their birth cohort delivered their last child at a slightly higher age and overall had slightly more children. This could be explained by a better maintenance of both reproductive health and overall health that supports longevity. Next, we studied whether the genetic component in familial longevity might associate with that of reproductive health, possibly pointing to common mechanisms in of both traits. However, females descending from such long-lived families do not have a different number of offspring or age at last child than females from non-long-lived families. Moreover, in the LLS, there was no evidence for an association between the genetic predisposition for a delayed age at menopause and familial longevity, as measured by a score indicating the proportion of long-lived ancestors. Hence, we conclude that a high age of last childbirth and number of offspring are markers of good reproductive health and overall health supporting longevity. They are not, however, explained by the genetic component in longevity, neither in the LINKS population nor in the Leiden Longevity study. Finally, the heritable component underlying the clinical extremes of age at menopause as represented by a PRS ^7^ does not appear to coincide with that in familial longevity.

Our study affirms previous research that supports a relationship between a later age at last childbirth and increased post-reproductive survival^2, 3, 15–19^ and the notion that the functioning of the reproductive system can be representative of females’ health, not necessarily driving it as our data imply. The ability for late reproduction and a larger number of offspring could be facilitated by a longer reproductive period, i.e. later age at menopause. Genes of DNA repair strongly relate to age at menopause^7^, suggesting that the latter is the result of overall somatic aging^20^. Although the genetic loci associated with age at menopause have not yet been directly correlated to human longevity or familial longevity, a meta-analysis relating SNPs to exceptional human longevity (in single cases) reported a correlation with several of the same SNPs that related to age at menopause^21^. Because the genetic predisposition for a late onset of menopause is not significantly associated to the familial component of human longevity in our study, the late reproducing females’ health might also be affected by other factors that influence lifespan in singletons (long lived persons without long lived family members) such as good environmental circumstances, healthy lifestyles, or favorable social factors.

Besides oocyte quantity, oocyte quality is a necessary factor for reproductive success and is thought to be a causal factor of age-related fertility decline^22^. As females age, oocyte competence decreases, leading to an increased risk of aneuploidy and miscarriage, in turn leading to decreased fecundity. Suggested pathways of oocyte quality decline include deterioration of the maintenance of mitochondrial function^23^ and the intrafollicular processes of DNA translation^24^. These processes are in turn thought to be subject to oxidative stress, to which the oocyte becomes more vulnerable with increasing age, as shown in animal models^25^. Fertility treatments such as in vitro fertilization (IVF) have previously, though not consistently, been linked to adverse cardiovascular outcomes in the short and long term^26–28^, which could suggest an adverse aging profile for the sub-fertile population, but it is not clear whether this can be attributed to effects of the treatment or population risk. While it is possible that the influence of the DNA damage repair genes associated with age at menopause additionally extends to oocyte quality, this remains to be further determined. Perhaps in oocytes, a group of cells that spend most of their life in senescence, the pathways for cell maintenance are regulated somewhat differently than in somatic cells. This could be another explanation for the lack of association between familial longevity and the polygenic risk score for menopause as well as reproductive outcomes in our study. It is also possible that fecundity, and thus oocyte quality, cannot be adequately measured through the proxies of age at childbirth and number of offspring. In addition, because we study a by-definition relatively healthy group of females who lived to age 50 and underwent at least one successful pregnancy and delivery, our study precludes an in-depth inquiry into the association between infertility or involuntary childlessness and longevity.

The method of linkage of families in the historical cohort makes our study uniquely suited to study the familial effects of reproduction and longevity. The methodological selection of the study population as well as the population size add to its strengths. Due to the historical nature of the data, the results are not influenced by the use of hormonal contraception or assisted reproductive techniques, therefore allowing for a reasonable assumption of unrestricted natural fertility.

Our results are limited by the obligatory use of proxy variables for fertility and reproductive success, as we were limited to the data stored in governmental registries. Furthermore, as mentioned we included a relatively healthy group of individuals who lived to be at least 50 years old. It is possible that this selection excludes individuals with accelerated aging genotypes and therefore attenuates any associations of reproduction with longevity.

In conclusion, we affirm that a late age at last childbirth is associated with a longer lifespan, and that traits of reproductive success seem to be markers of females’ health in middle age, likely acquired by good environmental circumstances. Furthermore, we conclude that neither parental nor more extended ancestral familial longevity are characterized by reproductive success.

## Data Availability

All data produced in the present study are available upon reasonable request to the authors

