## Supplementary information for "Late reproduction is associated with extended female survival but not with familial longevity"

**Supplementary Figure 1: Family-tree overview of the LINKS data structure**

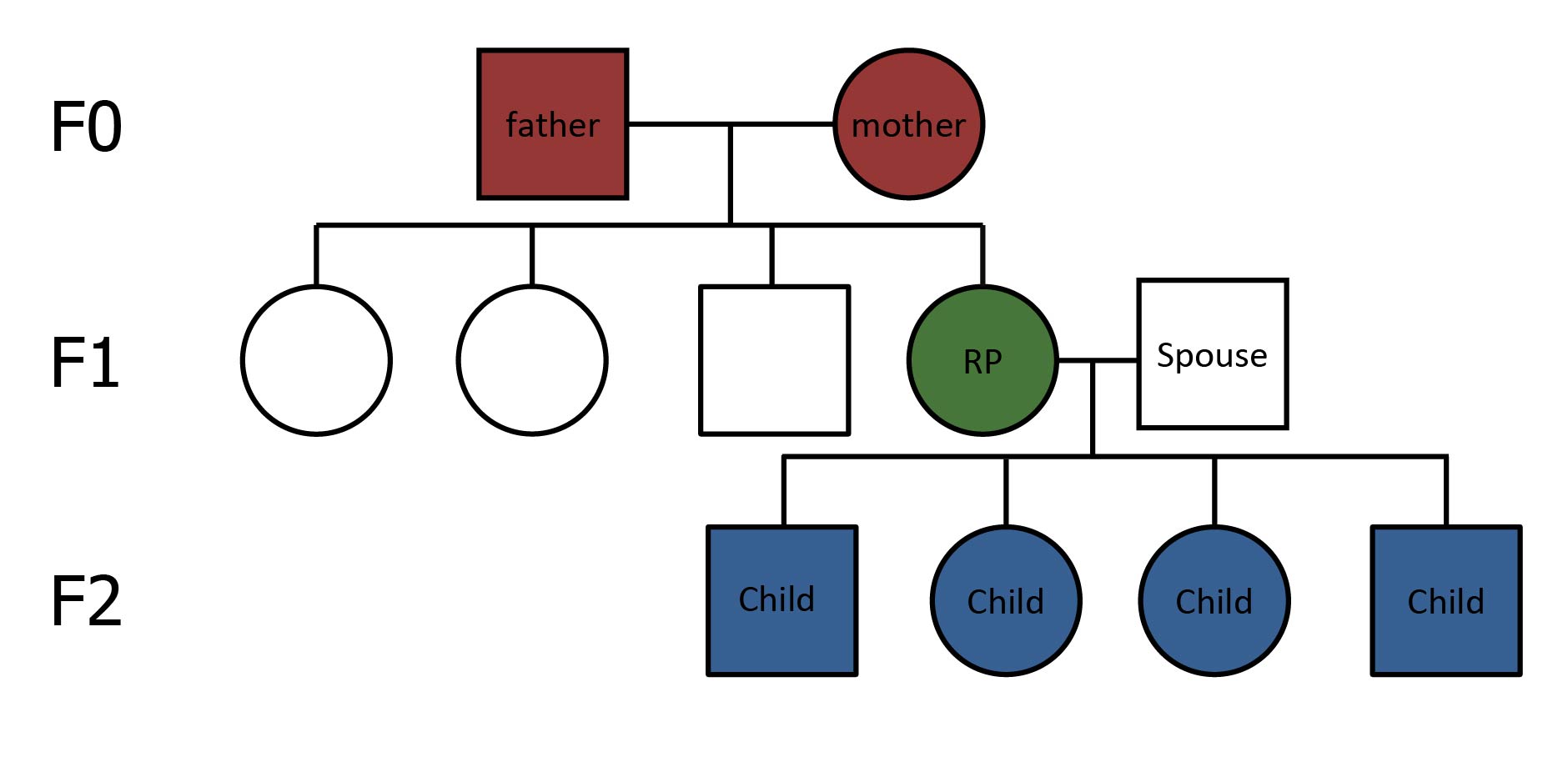

F indicates filial generation 0-2. Two generations were identified in the dataset; F0 and F1, of which the F1 generation is the index generation comprising the study participants. The F0 generation was selected by identifying couples who were married between 1812 and 1850 and had at least two children, ensuring that the F1 persons had at least one sibling. The families were mutually exclusive, meaning that a parent in the F0 generation could only contribute data for a single family. From the F1 generation, the LINKS research persons (RP) were selected. . Reproductive characteristics of the RPs are derived using information of their children (F3 generation). Rounds represent females and squares represent males. Note that in this example a single RP was indicated but if multiple female siblings met the inclusion criteria multiple siblings could be an RP.
